# The impact of exercise training on the brain and cognition in T2DM, and its physiological mediators: a systematic review

**DOI:** 10.1101/2024.09.19.24313875

**Authors:** Jitske Vandersmissen, Ilse Dewachter, Koen Cuypers, Dominique Hansen

## Abstract

**Background:** Type 2 diabetes (T2DM) affects brain structure and function, and is associated with an increased risk of dementia and mild cognitive impairment. It is known that exercise training has a beneficial effect on cognition and the brain, at least in healthy people, but the impact of exercise training on cognition and the brain remains to be fully elucidated in patients with T2DM.

**Methods:** This paper systematically reviews studies that evaluate the effect of exercise training on cognition in T2DM, and aims to indicate the most beneficial exercise modality for improving or preserving cognition in this patient group. In addition, the possible physiological mediators and targets involved in these improvements are narratively described in the second part of this review. Papers published up until the end of June 2024 were searched by means of the electronic database PubMed. Studies directly investigating the effect of any kind of exercise training on the brain or cognition in patients with T2DM, or animal models thereof, were included, with the exception of human studies assessing cognition only at one time point, and studies combining exercise training with other interventions (e.g. dietary changes, cognitive training, etc.).

**Results:** For the systematic part of the review, 24 papers were found to be eligible. 20 out of 24 papers (83.3%) showed a significant positive effect of exercise training on cognition in T2DM, of which four studies only showed a moderate significant effect. Four papers (16.7%) did not show a significant effect of exercise on cognition in T2DM, but two of them did show a positive trend. Similar effects were found for resistance and endurance exercise, with both possibly requiring a minimal intensity to reach cognitive improvement. In addition, BDNF, lactate, leptin, adiponectin, GSK3β, GLP-1, the AMPK/SIRT1 pathway, and the PI3K/Akt pathway were identified as plausible mediators directly from studies investigating the effect of exercise training on the brain in T2DM.

**Conclusion:** Overall, exercise training beneficially affects cognition and the brain in T2DM, with resistance and endurance exercise having similar effects. However, additional studies comparing the effect of different exercise intensities are needed to determine the optimal exercise intensity for cognitive improvement. Furthermore, we were able to define several mediators involved in the effect of exercise training on cognition in T2DM, but further research is necessary to unravel the entire process.

This review demonstrates a positive effect of exercise training on the brain in T2DM, but simultaneously emphasizes the need for additional studies on this topic. BDNF, lactate, leptin, adiponectin, GSK3β, GLP-1, the AMPK/SIRT1 pathway, and the PI3K/Akt pathway were identified as factors mediating the effect of exercise on the brain in T2DM.

## 1. Introduction

### 1.1 Cognitive decline in aging

Cognitive decline is inherent to aging. As we age, synaptic plasticity reduces, neural mitochondrial function declines, epigenetic changes occur, etc., all contributing to decreased cognitive abilities [1–5]. Especially fluid intelligence, defined as the ability to solve problems, process new information, and learn new things, declines with aging. Fluid intelligence consists of several domains, including executive function, psychomotor ability, memory, and processing speed [6, 7].

Aging is associated with a decline in both grey and white matter volume, accompanied by ventricular enlargement and cortical thinning, as well as a decline in brain function [7, 8]. Amyloid β plaques are not only found in Alzheimer’s disease (AD) patients, but also in 20-30% of healthy adults, and are speculated to contribute to this neuronal loss [6, 9]. Also neuronal volume and the number of neuronal connections decreases due to a decline in the complexity of dendrite arborization, decreased neuritic spines, and reduced dendrite length [6, 10]. In addition, demyelination and decreased white matter integrity contribute to functional impairment of the brain [8].

### 1.2 Cognitive dysfunction in T2DM

T2DM patients are prone to more severe cognitive decline than what occurs during normal aging. A large fraction (45%) of T2DM patients experience cognitive dysfunction and have an increased risk of dementia [11–16]. Subjects with diabetes have a relative risk of 1.46 for AD, and a relative risk of 1.51 for any form of dementia [17]. Especially executive functioning, memory, attention, and information processing speed have been shown to be affected in T2DM [14, 18, 19]. This cognitive impairment is related to the affected brain structure and metabolism in T2DM. Decreased regional grey matter volume as well as altered intrinsic activity in the default mode network have been demonstrated in these patients [20–22]. Multiple pathophysiologies have been suggested as the cause of this, including cardiovascular complications, chronic low-grade inflammation, and hyperglycemia [14, 15, 18].

Several studies have demonstrated a link between T2DM and AD, also called type 3 diabetes. These studies focus on the fact that insulin signalling also plays an important role in the brain, and that T2DM and AD seem to share a number of pathophysiological processes such as amyloid β plaques, disturbed cerebral glucose metabolism, tau hyperphosphorylation, and inflammation [18, 23–26]. Insulin and insulin-like-growth-factor (IGF)-1 are important for neuronal survival and brain function. Numerous insulin receptors (IR) and IGF-1 receptors (IGF-1R) can be found in the brain, with a large amount being present in the hippocampus. One of the major downstream pathways of the IR is the PI3K/Akt pathway, which plays an important role in the regulation of brain function, and in the inactivation of GSK3β, which is an enzyme able to phosphorylate tau at pathological tau epitopes [23, 27, 28]. Reduced cerebral IR activation and insulin levels have been demonstrated in AD [25, 29], highlighting similar processes in AD and T2DM. Moreover, insulin-degrading enzyme is capable of clearing amyloid β peptides that aggregate into amyloid plaques, a key pathological hallmark of AD. In T2DM, competitive binding of high insulin concentrations and amyloid β peptides with this enzyme could be involved in decreased amyloid β clearance, thereby potentially contributing to increased accumulation of amyloid β peptides, and formation of amyloid β plaques [15, 30, 31].

### 1.3 Positive effect of exercise on cognitive function

In healthy people, exercise has a positive effect on cognitive function [32–34]. This effect is mediated by multiple processes. Muscle contraction during exercise releases myokines, which stimulate the production of neurotrophic factors such as brain-derived neurotrophic factor (BDNF), promoting neurogenesis. Additionally, the exercise-induced release of anti-inflammatory factors contributes to balancing the brain’s redox status, counteracting many pathological processes [35–38]. Research shows that exercise reduces the age-related decrease in hippocampal volume [39–41], which, in combination with increased levels of neurotrophic factors, contributes to the maintenance of memory and neuroplasticity [36, 42, 43]. Moreover, there is a positive association between cardiorespiratory fitness levels and whole-brain and white matter volume in patients with early-stage AD [44]. In this way, exercise reduces the risk of dementia and neurodegenerative diseases [36, 45–47].

Lactate has been identified as an important contributor to the positive effect of exercise on cognitive function. During anaerobic exercise, lactate is released from contracting muscles, eventually leading to increased lactate levels in the brain after crossing the blood-brain-barrier (BBB). This results in an increased expression of brain plasticity genes such as BDNF, Arc, and c-fos, contributing to neurogenesis [48–50]. In addition, lactate also binds to hydroxycarboxylic acid receptor 1 (HCAR1), which leads to an increase in vascular endothelial growth factor (VEGF), stimulating angiogenesis and thereby contributing to increased cognitive function [50–52]. Also the myokine irisin has been shown to positively influence cognition. Synaptic plasticity and memory in amyloid pathology mimicking AD mouse models can be rescued by boosting brain levels of FNDC5/irisin, and peripheral overexpression of FNDC5/irisin rescues memory impairment [53]. Cathepsin b is another myokine which has been associated with improved cognition. It is known to play a role in both neurogenesis and angiogenesis [54, 55]. Cathepsin b levels increase in mouse and human plasma in response to exercise, which is positively correlated with memory, while cathepsin b-knockout mice do not show any cognitive improvements following running exercise [56]. This suggests that cathepsin b is mandatory for the positive effect of exercise on cognition.

### 1.4 The association of physical activity with cognitive function in T2DM

Because of the above-mentioned positive effects of exercise on cognitive function, and to get more insight into whether this translates to T2DM, some studies have explored the association between physical activity (PA) and cognition in T2DM.

For example, one study found significantly higher cognitive scores in an active group of T2DM patients compared to a sedentary group of T2DM patients [57]. They also found a significant negative correlation (r = −0.2, P = 0.03) between cognition and BMI in the sedentary group, and a significant positive correlation (r = 0.55, P = 0.01) between cognition and minutes of weekly exercise in the active group. Another study found that active elderly T2DM patients had a significantly slower rate of cognitive decline compared to non-active/sedentary elderly T2DM patients, more specifically in the global cognition (p = 0.005), executive functioning (p = 0.014), and attention/working memory (p = 0.01) domains [58]. These findings were confirmed by multiple other studies [59–61].

In a study investigating the influence of the BDNF Val66Met polymorphism on the association between cognition and PA in diabetic patients, it was found that carriers of the Met-allele showed significantly higher scores of words recall (p < 0.001), mental status (p = 0.004), and total cognition, (p = 0.04), and had a significantly higher education level. Overall, PA was associated with better total cognition, words recall, and mental status, no matter the intensity. This association was strongest in the Met/Met carrier group, and stronger for females than for males within this group. They also found that light to moderate PA showed greater association with cognitive domains than moderate to vigorous PA. These findings thus suggest that female Met/Met diabetic carriers cognitively benefit the most from PA, and that light to moderate PA has the most influence on the brain [61].

Another study examined the individual and joint influence of diabetes status, apolipoprotein E (APOE) ε4, and PA on the risk of dementia and cognitive impairment without dementia (CIND) in cognitively normal older adults. The risk of dementia and CIND was higher in diabetic patients and APOE ε4 carriers who reported low levels of moderate to vigorous PA. The risk was even found to be nearly 10-fold higher for physically inactive diabetic APOE ε4 carriers. Higher levels of PA were associated with lower dementia/CIND risk [60].

Overall, these findings suggest that higher levels of PA are associated with better cognitive function and less cognitive decline in T2DM. However, based on these studies alone, one cannot assume causality. In addition, there is a possibility of reverse causality, where reduced cognitive function contributes to lower levels of PA. Therefore, several studies have investigated the acute effect of exercise on cognitive function in T2DM.

### 1.5 The acute effect of exercise on cognitive function in T2DM

The acute effect of exercise on cognition in T2DM has mainly been investigated in the domain of executive function.

One study found that both endurance and resistance training acutely improved inhibitory control and response time in T2DM patients [62]. In addition, moderate-intensity integrated concurrent exercise (ICE), consisting of both endurance and resistance training, acutely improved all three aspects of executive function (inhibition, conversion and refresh function) in cognitively normal hospitalised T2DM patients, while resistance exercise improved inhibition, and endurance exercise only caused significant improvements in the refresh function. The ICE-induced improvements in executive function were accompanied by a simultaneous increase in cerebral blood flow in the dorsolateral prefrontal cortex (DLPFC), the frontal pursuit area (FPA), and the orbitofrontal cortex (OFC), while resistance exercise showed a corresponding activation of DLPFC and FPA, and endurance exercise increased cerebral perfusion in the FPA, OFC and Broca region [63]. These results suggest that an exercise-induced increase in cerebral blood flow in certain areas can elicit specific cognitive improvements. This knowledge can be used to customise exercise programs in T2DM patients in order to obtain the desired effect.

These studies already give some insight into the effect of exercise on cognition in T2DM. However, with the aim to incorporate exercise in T2DM treatment for cognitive improvement, it is necessary to know the effect of *long-term exercise training* on the brain, which can be different from the acute effect. Therefore, this review will focus on pre-post intervention studies. This review only describes RCTs investigating the effect of exercise training on cognition and brain structure in T2DM and animal models thereof, with special emphasis on the mediators and targets involved in this. So far, this is the first review of this kind. The knowledge provided by this review could more reliably provide insights in the mechanisms of cognitive decline and cognitive improvement in T2DM, and could be used to design more specific exercise programs to target the brain in T2DM patients.

## 2. Methods

This review consists of two parts, the first of which is a systematic review of the papers investigating the effect of exercise training on cognition in T2DM. The second part narratively describes studies that show beneficial brain changes or brain changes in combination with cognitive improvement in T2DM as a result of exercise training, and aims to identify the involved mediators by pooling studies with similar findings.

### 2.1 Search methods

The papers described in the systematic part of this review were found by entering the following search string into PubMed: (exercise OR “physical activity” OR “aerobic exercise” OR “aerobic training” OR “resistance exercise” OR “resistance training” OR “combined exercise” OR “combined training” OR “endurance exercise” OR “endurance training”) AND (T2DM OR diabetes OR “type 2 diabetes”) AND (memory OR “executive function*” OR “processing speed” OR attention OR brain OR cognition OR “cognitive function”). The filters “randomised controlled trial” and “clinical trial” were applied. Relevant related articles suggested by PubMed when accessing articles found by entering the aforementioned search string were also considered. Reference lists of reviews in line with the present one [64–72] were examined as well. The current review discusses articles published up until the end of June 2024. Duplicate articles and studies of poor quality were removed (Fig. 1). For the narrative part of the review, only studies directly investigating the effect of exercise on the brain in T2DM were considered to identify plausible mediators and pathways.

**Fig. 1.**
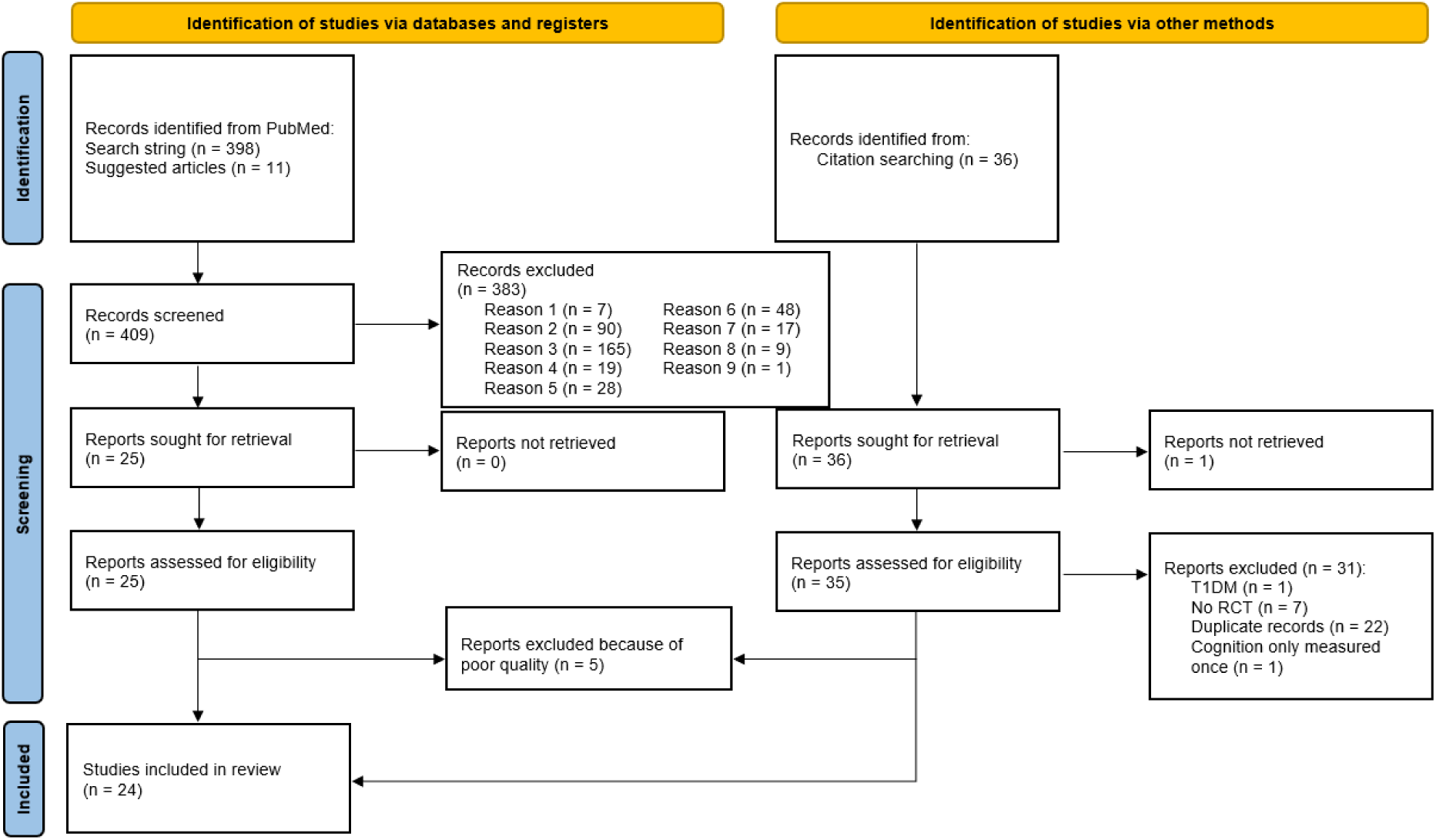
Prisma flow chart of study selection procedure. Reason 1: Does not investigate the effect of exercise training; Reason 2: Does not investigate the effect of exercise training on cognition; Reason 3: Does not investigate the effect of exercise training on cognition in T2DM; Reason 4: Not conducted in T2DM; Reason 5: Does not investigate the effect on cognition; Reason 6: Does not investigate the effect on cognition in T2DM; Reason 7: Does not investigate the effect of exercise training in T2DM; Reason 8: Does not investigate the effect of exercise training on its own (diet, cognitive training,…); Reason 9: Article not available in English. RCT = randomised controlled trial; T1DM = type 1 diabetes. From: Page MJ, McKenzie JE, Bossuyt PM, Boutron I, Hoffmann TC, Mulrow CD, et al. The PRISMA 2020 statement: an updated guideline for reporting systematic reviews. BMJ 2021;372:n71. doi: 10.1136/bmj.n71.

**Fig. 2.**
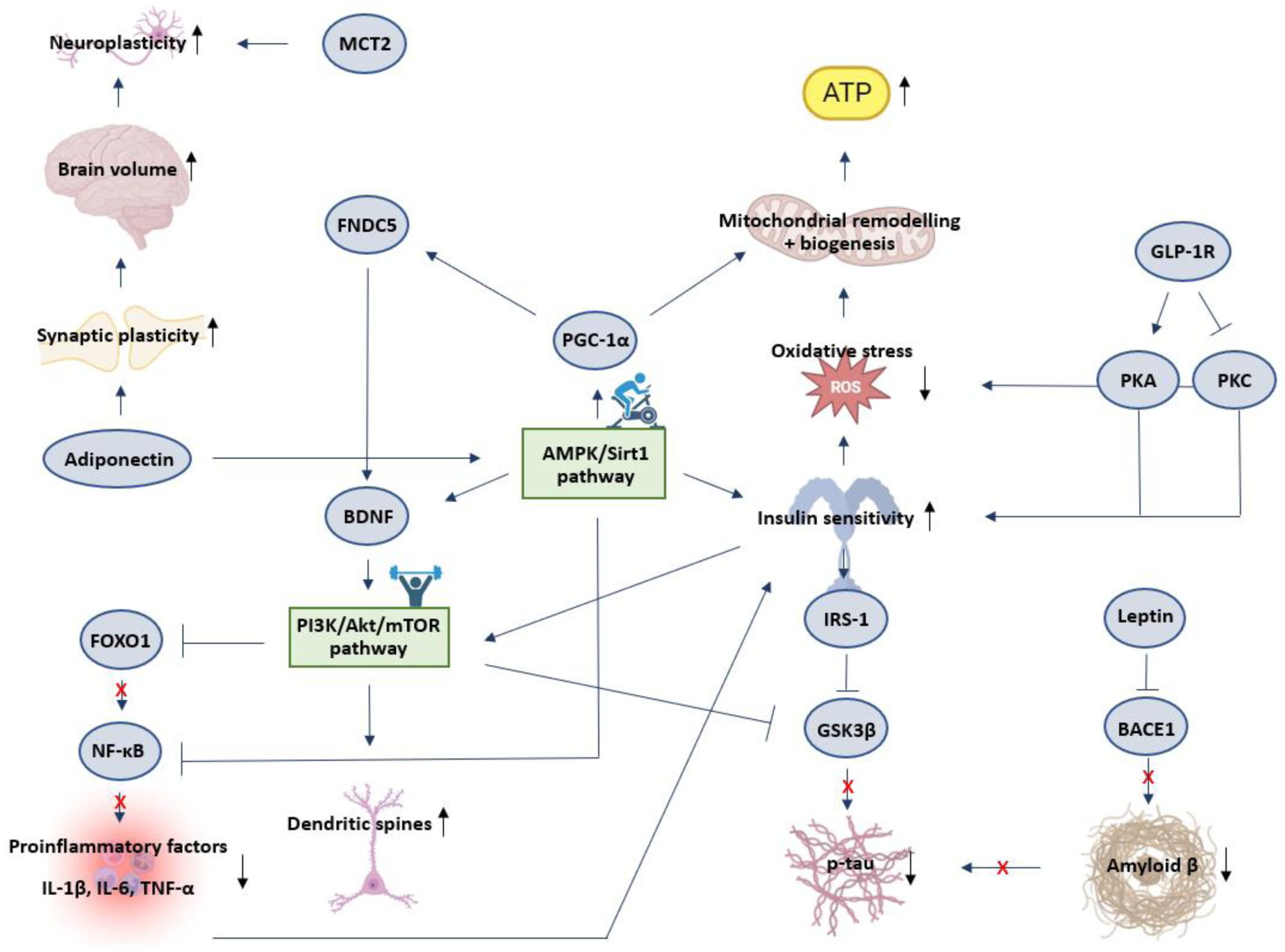
Schematic representation of mediators and targets involved in the effect of exercise training on the brain in T2DM. Sharp blue arrows indicate stimulation or increase. Blunt blue arrows indicate inhibition. Aβ = amyloid β; AMPK = AMP-activated Protein Kinase; ATP = Adenosine Triphosphate; BDNF = Brain-Derived Neurotrophic Factor; FNDC5 = Fibronectin type III Domain-Containing protein 5; FOXO1 = Forkhead box protein O1; GLP-1R = Glucagon-Like Peptide 1 Receptor; GSK3β = Glycogen Synthase Kinase-3 beta; IRS-1 = Insulin Receptor Substrate 1; IL-1β = Interleukin 1beta; IL-6 = Interleukin 6; MCT2 = Monocarboxylate Transporter 2; mTOR = mammalian Target Of Rapamycin; Nf-κB = Nuclear Factor kappa-light-chain-enhancer of activated B-cells; PGC-1α = Peroxisome proliferator-activated receptor Gamma Coactivator 1-alpha; PI3K = Phosphoinositide 3-Kinase; PKA = Protein Kinase A; PKC = Protein Kinase C; p-tau = phosphorylated tau; ROS = Reactive Oxygen Species; Sirt1 = Sirtuin 1; TNF-α = Tumor Necrosis Factor alpha.

### 2.2 Eligibility

The PICO (population, intervention, comparison, outcome) method was used to determine whether articles were eligible for inclusion in this review. In order to be included in the systematic part of this review, the articles had to be written in English, and had to investigate the effect of any kind of exercise training on cognition in T2DM patients or T2DM animal models. Articles discussing only type 1 diabetes were excluded, as well as articles only investigating depression, anxiety, or other mental disorders in T2DM. Also studies investigating the effect of exercise training in combination with another intervention (e.g. dietary changes, cognitive training, etc.) were excluded. Studies in which cognitive function was only assessed at one time point in human subjects were excluded since these make it impossible to accurately assess the effect of exercise training on cognition (pre- and post-intervention assessments mandatory).

### 2.3 Quality control

Quality control of the included human studies was conducted by means of the PEDro scale, which assesses internal validity [73]. With this tool, a maximal score of 10 can be reached, indicating optimal methodological quality. However, it was impossible for the majority of the studies included in this review to reach a maximal score of 10, since in exercise interventions it is not feasible to blind the participants and the therapists who administered the therapy. Because of this, a score of >4/10 instead of >6/10 was required for inclusion. Quality of the animal studies was assessed by means of the Collaborative Approach to Meta-Analysis and Review of Animal Data from Experimental Studies (CAMARADES) checklist [74]. Animal studies with a score of <6/10 were excluded. If nothing was mentioned about the criterion, it was indicated as not fulfilled.

**Table 1.** PEDro characteristics of included human studies.

|  | 1 | 2 | 3 | 4 | 5 | 6 | 7 | 8 | 9 | 10 | 11 | Total |
| --- | --- | --- | --- | --- | --- | --- | --- | --- | --- | --- | --- | --- |
| Leischik et al. (2021) | ✓ | ✓ | ✓ | ✓ | X | X | X | ✓ | ✓ | ✓ | ✓ | 7/10 |
| Anderson-Hanley et al. (2012) | ✓ | ✓ | X | ✓ | X | X | X | X | ✓ | ✓ | ✓ | 5/10 |
| Ploydang et al. (2023) | ✓ | ✓ | X | ✓ | X | X | X | ✓ | ✓ | ✓ | ✓ | 6/10 |
| Wang et al. (2023) | ✓ | ✓ | ✓ | ✓ | X | X | ✓ | X | ✓ | ✓ | ✓ | 7/10 |
| Zhao et al. (2022) | ✓ | ✓ | ✓ | X | ✓ | ✓ | ✓ | ✓ | ✓ | ✓ | ✓ | 9/10 |
| Yamamoto et al. (2021) | ✓ | ✓ | X | ✓ | X | X | X | ✓ | ✓ | ✓ | ✓ | 6/10 |
| Teixeira et al. (2019) | ✓ | ✓ | X | ✓ | X | X | X | ✓ | ✓ | X | ✓ | 5/10 |
| Silveira-Rodrigues et al. (2021) | ✓ | X | X | ✓ | X | X | X | ✓ | ✓ | ✓ | ✓ | 5/10 |
| Espeland et al. (2017) | ✓ | ✓ | X | ✓ | X | ✓ | ✓ | ✓ | ✓ | ✓ | ✓ | 8/10 |
| Silveira-Rodrigues et al. (2023) | ✓ | X | X | ✓ | X | X | X | ✓ | ✓ | ✓ | ✓ | 5/10 |
| Ghodrati et al. (2023) | ✓ | ✓ | X | ✓ | X | X | X | ✓ | ✓ | ✓ | ✓ | 6/10 |
| Martinez-Velilla et al. (2021) | ✓ | ✓ | X | ✓ | X | X | ✓ | ✓ | ✓ | ✓ | ✓ | 7/10 |
| Callisaya et al. (2017) | ✓ | ✓ | ✓ | ✓ | ✓ | X | ✓ | ✓ | ✓ | ✓ | ✓ | 8/10 |
| Ghahfarrokhi et al. (2024) | ✓ | ✓ | ✓ | ✓ | X | X | ✓ | ✓ | ✓ | ✓ | ✓ | 8/10 |
| Shellington et al. (2018) | ✓ | ✓ | ✓ | ✓ | X | X | X | X | ✓ | ✓ | ✓ | 5/10 |
| Chen et al. (2023) | ✓ | ✓ | X | ✓ | X | X | ✓ | ✓ | ✓ | ✓ | ✓ | 7/10 |
| Cai et al. (2019) | ✓ | X | X | ✓ | X | X | X | ✓ | ✓ | ✓ | ✓ | 5/10 |
| Coelho Junior et al. (2018) | ✓ | X | X | ✓ | X | X | X | ✓ | ✓ | ✓ | ✓ | 5/10 |

✓ = fulfilled, X = not fulfilled, 1 = Eligibility criteria specified, 2 = Randomisation, 3 = Allocation concealed, 4 = Groups similar at baseline, 5 = Blinding of subjects, 6 = blinding of therapists, 7 = Blinding of assessors, 8 = Measures obtained for more than 85% of initial subjects, 9 = Intention-to-treat analysis, 10 = Between-group statistical comparisons, 11 = point measures and measures of variability

**Table 2.** CAMARADES characteristics of included animal studies.

|  | 1 | 2 | 3 | 4 | 5 | 6 | 7 | 8 | 9 | 10 | Total |
| --- | --- | --- | --- | --- | --- | --- | --- | --- | --- | --- | --- |
| Parsa et al. (2021) | ✓ | ✓ | ✓ | X | ✓ | ✓ | ✓ | X | ✓ | ✓ | 8/10 |
| Shekarchian et al. (2023) | ✓ | ✓ | ✓ | X | X | ✓ | ✓ | X | X | ✓ | 6/10 |
| Jesmin et al. (2022) | ✓ | ✓ | ✓ | X | X | ✓ | ✓ | X | X | ✓ | 6/10 |
| Lang et al. (2020) | ✓ | ✓ | ✓ | ✓ | ✓ | ✓ | ✓ | X | ✓ | ✓ | 9/10 |
| Shima et al. (2017) | ✓ | ✓ | ✓ | X | X | ✓ | ✓ | X | ✓ | ✓ | 7/10 |
| Shima et al. (2023) | ✓ | ✓ | X | X | X | ✓ | ✓ | X | ✓ | ✓ | 6/10 |

✓ = fulfilled, X = not fulfilled, 1 = Published in peer-reviewed journal, 2 = Control of temperature, 3 = Randomisation, 4 = Allocation concealed, 5 = Blinding of assessors, 6 = No anaesthetics with marked intrinsic properties, 7 = Use of animals with diabetes, 8 = Sample size calculation, 9 = Compliance with regulatory requirements, 10 = Statement regarding conflict of interest

## 3. Results

### 3.1 Systematic review on the effect of exercise training on cognition in T2DM

#### 3.1.1 Endurance exercise training

##### Human studies

Endurance exercise as simple as walking can already significantly improve cognition in T2DM patients. In a study by Leischik et al. (2021), T2DM patients were randomised into a walking group, pedometer group or control group. The walking group had to walk for 40 min, 3 times a week. The pedometer group had to reach 10 000 steps per day. Both the walking group and the pedometer group, the latter of which managed to reach an average of 8700 steps per day, showed significant improvements in attention and non-verbal memory. Verbal memory also significantly improved in the walking group [75]. Besides normal walking, also aquatic Nordic walking performed for 60 min 3 times a week for 12 weeks, has proven to cause significantly increased Montreal Cognitive Assessment (MoCA) scores in T2DM patients. However, no significant improvements were seen in the MMSE, trail making test part B, or Stroop colour and word test [76].

Multiple other studies have demonstrated the positive effect of endurance exercise training on cognition in T2DM. One study found that 6 months of endurance exercise significantly improves executive function in older adults with glucose intolerance, including prediabetic and newly diagnosed diabetic patients. Memory, however, showed no significant improvements [77]. Another study investigated the relationship between 3 months of endurance exercise and executive function in 10 T2DM patients and 10 age-matched controls. Executive function was tested at three different time points: a pre-test to familiarise the participants with the tests, and to avoid practice effects in the post-exercise test, a pre-exercise test one month after the pre-test, and a post-exercise test. Differences between the pre-exercise and post-exercise test were assessed. A significantly improved performance was observed in the diabetic group for the Colour Trails 2 test, in which the participants needed to connect numbered dots in numerical order while alternating the colour of the dots. The time to complete the task decreased from 117 seconds to 92 seconds in the diabetic group, as opposed to a non-significant decrease from 103 seconds to 99 seconds in the healthy control group. No significant improvements were found for the Digit Span Backwards or Stroop C tests [78]. Additionally, in a study by Wang et al. (2023), the effect of endurance training on hippocampal volume and cognition was tested in 82 T2DM patients. The patients were divided into a control group and a training group. After one year of endurance training, they found significantly increased MMSE and MoCA scores in the training group compared to the control group, suggesting that endurance exercise can improve cognitive function in T2DM [79].

##### Animal studies

Also animal studies show a predominantly positive effect of endurance exercise training on cognition. Shekarchian et al. (2023) found that streptozotocin- and high fat diet-induced diabetic C57BL/6 J mice that received 4 weeks of swimming training scored significantly better on tests for working, spatial, and recognition memory compared to non-exercising diabetic mice [80]. Another study showed that swimming training for 12 weeks in streptozotocin-nicotinamide-induced type 2 diabetic rats improved exploratory behaviour, locomotor activity, passive avoidance memory, and non-spatial cognitive memory compared to their sedentary counterparts. However, this improvement was found to be insignificant [81].

The Morris water maze test has oftentimes been used to demonstrate the effect of endurance exercise on cognition in animal models of T2DM. In this test, the animal is placed in a circular swimming pool where a platform is hidden somewhere under the water surface. After a few practice rounds, the time it takes the animal to find the platform, and the length of the path that is followed, are measured. These measures give an indication of spatial learning and memory [82]. For example, one study looked into the effect of 4 months of both light and moderate intensity treadmill running on memory in pre-symptomatic Otsuka-Long-Evans-Tokushima fatty (OLETF) rats. Before the exercise intervention, the OLETF rats spent considerably less time in the quadrant area where the platform was located during the learning phase, compared to control Long-Evans Tokushima (LETO) rats. After the intervention, both exercise intensities showed to have improved memory function in the OLETF rats, since both escape latency and swim length were shortened [83]. Lang et al. (2020) also used the Morris water maze test to test the effect of 8 weeks of moderate-intensity treadmill exercise on memory in T2DM mice. The exercising T2DM mice showed a significantly reduced escape latency on day 4-5 compared to the non-exercising T2DM mice, and their number of platform crossings significantly increased over time [84]. Another similar study was performed where the effect of 4 weeks of moderate-intensity treadmill running in OLETF rats was investigated. They found that both swim path length and escape latency of the exercising OLETF rats were reduced at trials on days 2-4, and that their time spent in the platform area significantly improved after the exercise intervention [85]. Another study by Shima et al. (2023) assessed the effect of 4 weeks of light-intensity running on a forced exercise wheel bed in ob/ob mice. However, here they did not find an effect of exercise on the swim distance, escape latency, or speed. They did find that the times of crossing the target platform during the probe test in the exercised ob/ob mice did not differ significantly from that in the control C57BL/6 mice, and that it was greater than in the sedentary ob/ob mice, but not significantly [86].

Based on these studies, endurance exercise conducted for 40-60 min, 3-5 times per week enables cognitive improvements in T2DM patients. Especially memory shows to be sensitive to endurance exercise-induced improvement. However, two out of five human studies discussed here only found significant improvements in MoCA and/or MMSE scores. Since the MMSE and MoCA were designed as screening tools to diagnose mild cognitive impairment (MCI) [87, 88], they cannot be considered very reliable for detecting changes in cognitive function.

#### 3.1.2 Resistance exercise training

##### Human studies

The beneficial effect of resistance exercise on cognition in T2DM patients was shown by Zhao et al. (2022). They examined the effect of 12 months of power training on executive function, attention/speed, memory, and global cognition in older adults with T2DM, and aimed to determine whether there is an association between cognitive improvements and improvements in muscle strength, body composition, and/or endurance. 103 patients were divided into a power training group or a sham low-intensity group that performed the same exercises as the power training group, but without added weight. Cognition of both groups improved over time, with increased scores in the Trails A, Trails B, word list recall, and word list memory. In addition, improved memory was associated with both increased skeletal muscle mass and reduced body fat mass, and improvement in Trails B minus A in the power training group was associated with increases in knee extension strength [89]. Yamamoto et al. (2021) failed to find similar results. In their study, 60 T2DM patients aged 72.9 ± 2.4 years were divided into a control group, a resistance training group, and a resistance training group with leucine (an amino acid promoting muscle synthesis) supplementation. The training groups performed daily bodyweight resistance exercises and exercises with elastic bands every day at home, for a total period of 48 weeks. Cognitive function was assessed by means of the MMSE at baseline and after the intervention period. The MMSE score in the control group was significantly decreased to 27.5 ± 2.6 after 48 weeks, while the score in the training groups had not significantly changed. This caused the MMSE score at 48 weeks to be significantly higher in the training groups than in the control group [90].

Although both studies suggest a positive effect of resistance exercise training on cognition in T2DM, a sufficient number of studies to draw a definite conclusion is lacking. Moreover, one study only assessed cognitive function by means of the MMSE, limiting the information on cognitive function, and thus the reliability of the results.

#### 3.1.3 Endurance vs. resistance exercise training

##### Human studies

To determine the exercise modality most effective in improving cognition in T2DM, some studies compared the effect of endurance and resistance exercise training. Teixeira et al. (2029) investigated the effect of either endurance or resistance exercise on cognitive function in T2DM patients or patients with arterial hypertension. The patients were randomised into a resistance exercise group or an endurance exercise group. Both groups exercised at moderate intensity for 12 weeks. Cognitive function was assessed before and after the intervention by means of the mental test and training system (MTTS), consisting of the cognitrone (attention and concentration), the determination test (reaction time), and the visual pursuit test (selective attention). The group with both T2DM and hypertension, but not the group with only hypertension, showed a significantly improved performance in the cognitrone, but no significantly improved reaction time. No differences were observed between the endurance and resistance exercise group. These results show that both endurance and resistance exercise are capable of increasing attention and concentration in T2DM patients with arterial hypertension [91].

This study shows that endurance and resistance exercise training have a comparable positive effect on cognitive function in T2DM, however, additional evidence confirming these results is needed.

#### 3.1.4 Combined endurance and resistance exercise training

##### Human studies

Several studies have explored the effect of combined exercise training, consisting of both resistance and endurance exercise, sometimes in combination with flexibility or balance exercises, on cognition in T2DM patients. One study found significantly improved inhibitory control, working memory, cognitive flexibility, and attention after 8 weeks of combined exercise training in T2DM patients, compared to a control group [92]. The same research team also examined the effect of 8 weeks of combined exercise on plasma BDNF levels, executive function and long-term memory in T2DM patients. They observed significantly improved executive function following the combined exercise, while BDNF levels were not found to be significantly changed [93]. Espeland et al. (2027) conducted an exploratory analysis of data from the Lifestyle Interventions and Independence for Elders (LIFE) trial, which was a randomised controlled clinical trial of exercise intervention consisting of walking, resistance training, and flexibility exercises in sedentary non-demented T2DM patients and healthy subjects. Cognitive function was tested at baseline and 2 years after randomisation. They found that cognitive function, more specifically global cognitive function and delayed memory, significantly improved in the diabetic participants of the intervention group only, suggesting a beneficial effect of combined training on cognition in T2DM [94]. Additionally, one study determined the effect of 12 weeks of combined exercise training, consisting of endurance, resistance, and balance exercises, on cognition in women with T2DM. Cognition was tested by means of the MoCA, the digit symbol substitution test, and the forward digit span test. After the 12-week intervention, the exercise group scored significantly higher on the MoCA compared to the control group, and, with an increase of 3.1, improved significantly compared to baseline. However, neither of the groups showed improvement in the other tests [95].

In a study by Martinez-Velilla et al. (2021), 103 acutely hospitalised elderly T2DM patients were randomised to an exercise group or a control group. Cognitive function was assessed by means of the MMSE at baseline and at discharge. The exercise intervention consisted of a combination of resistance, balance, and walking exercises. The median length of stay, and thus of the intervention, was 8 days. There was no difference in MMSE score between both groups at baseline. However, after the intervention, the exercise group scored on average 1.6 points higher than the control group (23.7 vs. 22.1), which was found to be significant [96]. In addition, a pilot study looked into the effect of 6 months of a progressive endurance- and resistance-training program on the brain and cognition in T2DM. 50 T2DM patients were randomised into an intervention group and a control group. The intervention group performed 6 months of endurance and progressive resistance training. The control group received upper and lower limb stretching of light intensity and a gentle movement program, which were performed in the same volume, frequency and setting as in the intervention group. The intervention group showed improved hippocampal and total brain volumes, improved white matter integrity, and less decline in white matter volume. They also showed a better global cognitive score, and better performance on the Digit Symbol Coding Test, Rey Complex Copy test, Stroop C-D, Trail Making Test A and B, Hopkins verbal learning test (intermediate and recognition scores), and Controlled Oral Word Association Test compared to the control group [97]. Another pilot study examined the effect of 6 weeks of high-intensity low-volume (HIFT) vs. low-intensity high-volume (LIFT) functional training on cognition in cognitively impaired elderly T2DM patients. The HIFT group exercised three times a week at 100-120% of the lactate threshold, while the LIFT group exercised five times a week at 70-75% of the lactate threshold. The MMSE was used to diagnose cognitive impairment (MMSE ≤ 23). Processing speed, learning, memory, and attention were assessed by means of the Symbol Digit Modalities Test (SDMT), California Verbal Learning Test Second Edition (CVLT-II), Brief Visuospatial Memory Test-Revised (BVMT-R), and Stroop tests respectively. After the intervention, MMSE, Stroop, SDMT, CVLT-II and BVMT-R scores had improved significantly in the HIFT group, while only MMSE and Stroop scores, had improved significantly in the LIFT group. However, the only cognitive score change that was significantly different from the control group that did not receive an exercise intervention, was the change in Stroop scores in the HIFT group [98].

Overall, most studies suggest a positive effect of combined exercise training on cognitive function in T2DM. However, mixed findings indicate the need for additional studies.

#### 3.1.5 Other types of exercise

##### Human studies

Besides the well-known exercise forms such as running and cycling, some studies have explored the effect of unconventional exercise training on cognition in T2DM.

For example, one study compared the effect of 36 weeks of Tai Chi Chuan, a mind-body exercise, and 36 weeks of fitness walking on global cognitive function in T2DM patients. Both interventions significantly increased MoCA scores, with a significantly larger effect of Tai Chi Chuan compared to fitness walking [99].

The effect of low-intensity Qigong exercise on cognitive function in older T2DM patients was also investigated [100]. Qigong is an ancient Chinese exercise for mind-body integration and is often used in the prevention and treatment of chronic metabolic diseases [101]. The participants in the exercise group practiced Kinect-based Kaimai-style Qigong for 12 weeks. Based on MMSE scores at baseline and after the intervention, the Qigong group showed significant improvements in cognitive function compared to the control group [100].

Coelho Junior et al. (2018) investigated whether multicomponent exercise (MCE) improves executive function in T2DM. 72 T2DM patients performed MCE for 26 weeks. MCE consisted of 12 different exercises mimicking activities of daily live gestures, performed for 1 minute each, with each exercise immediately followed by a 2 minute walk. Although the patients showed an increased mobility after the intervention, they did not show improvements in executive function [102].

Finally, the effect of a 24-week square-stepping exercise (SSE) program on cognition in T2DM patients older than 49 years was also investigated. The researchers assessed global cognitive function, delineated into 4 domains: memory, reasoning, concentration and planning. The SSE group showed greater improvement in planning scores compared to the control group. However, the improvement in this specific domain might be related to the nature of the training program, where the participants are required to memorise patterns [103].

These findings suggest that unconventional exercise, often involving the mind to a larger extent than conventional exercise, can contribute to improved cognition in T2DM.

**Table 3.**
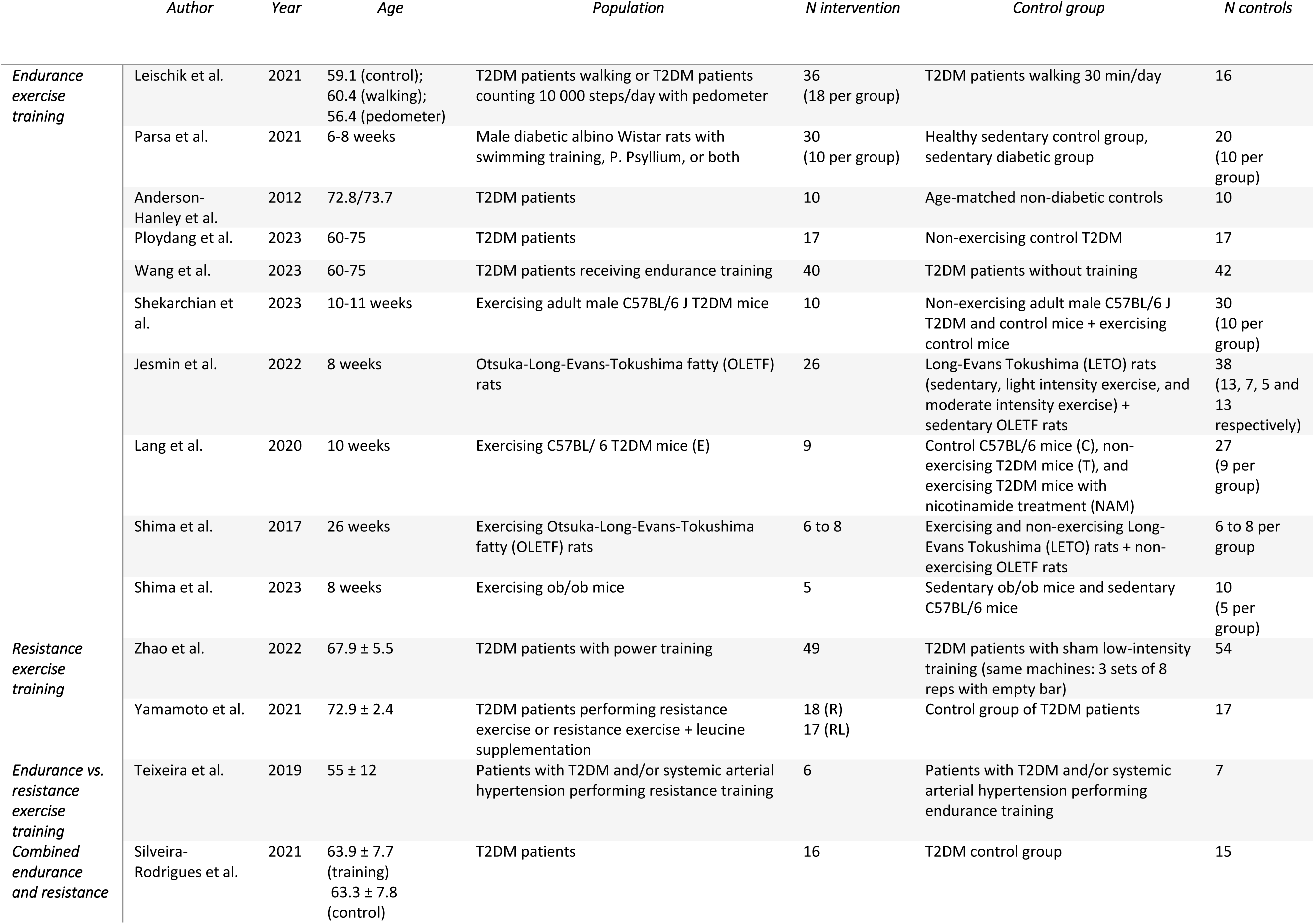
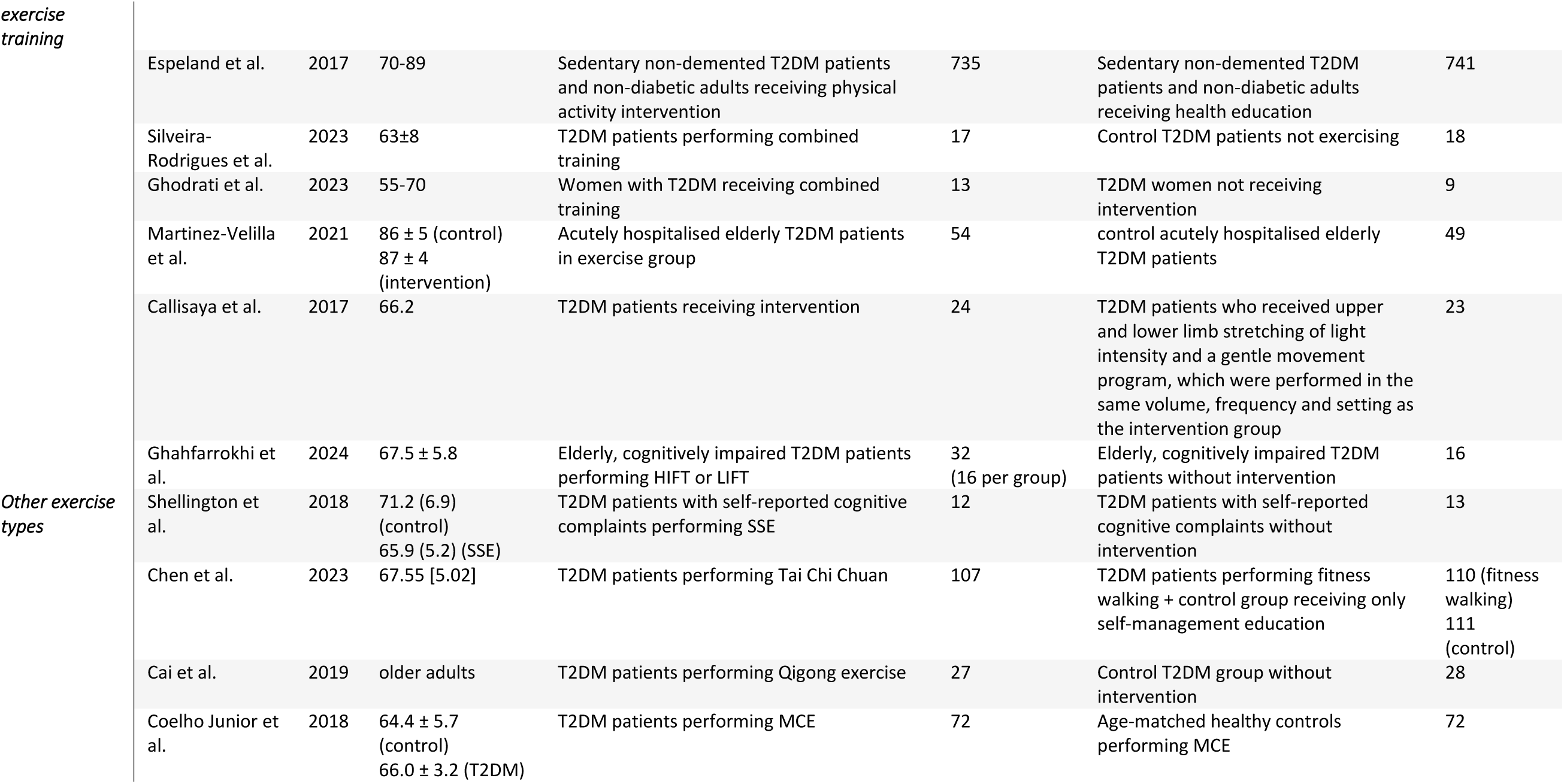
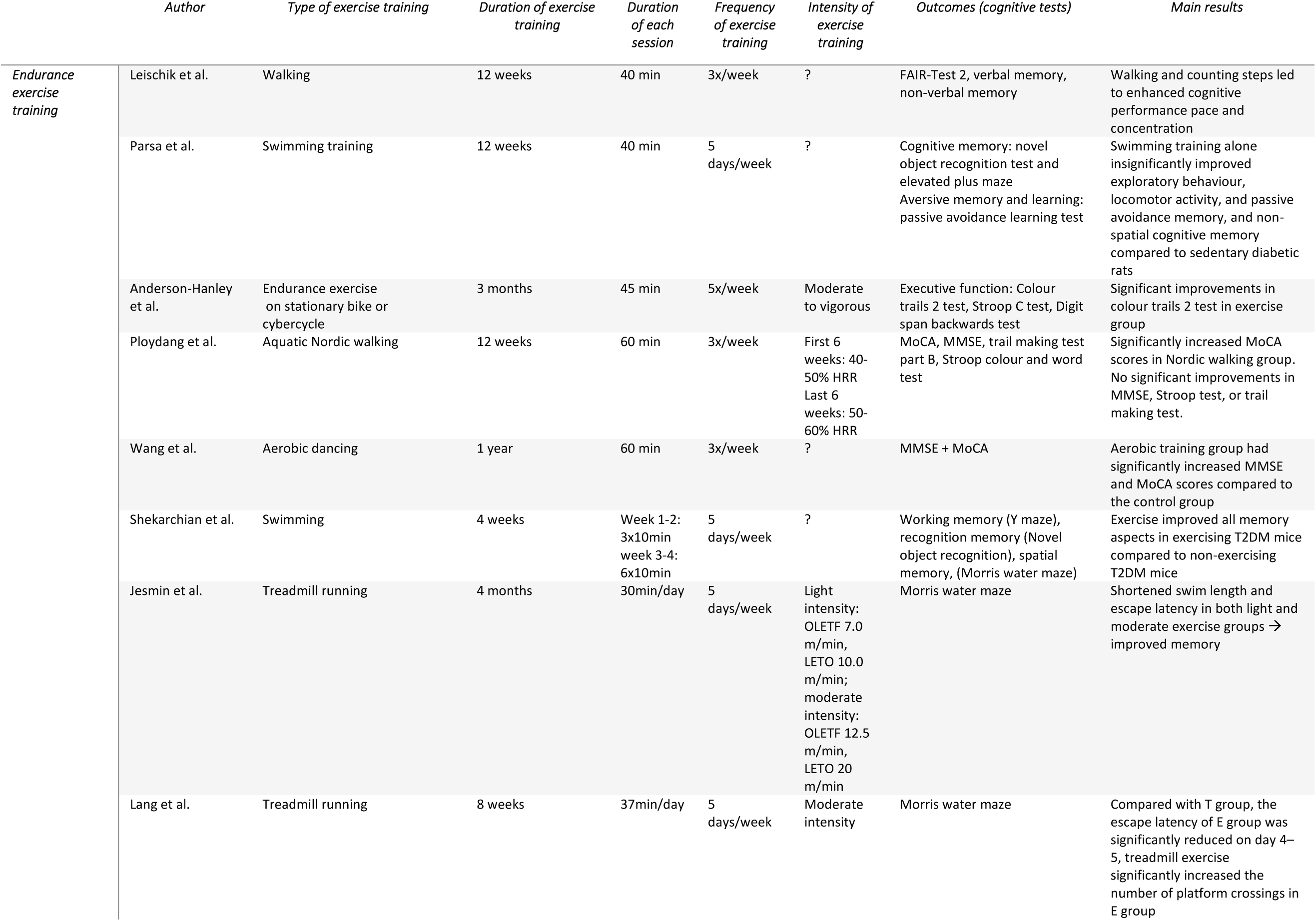
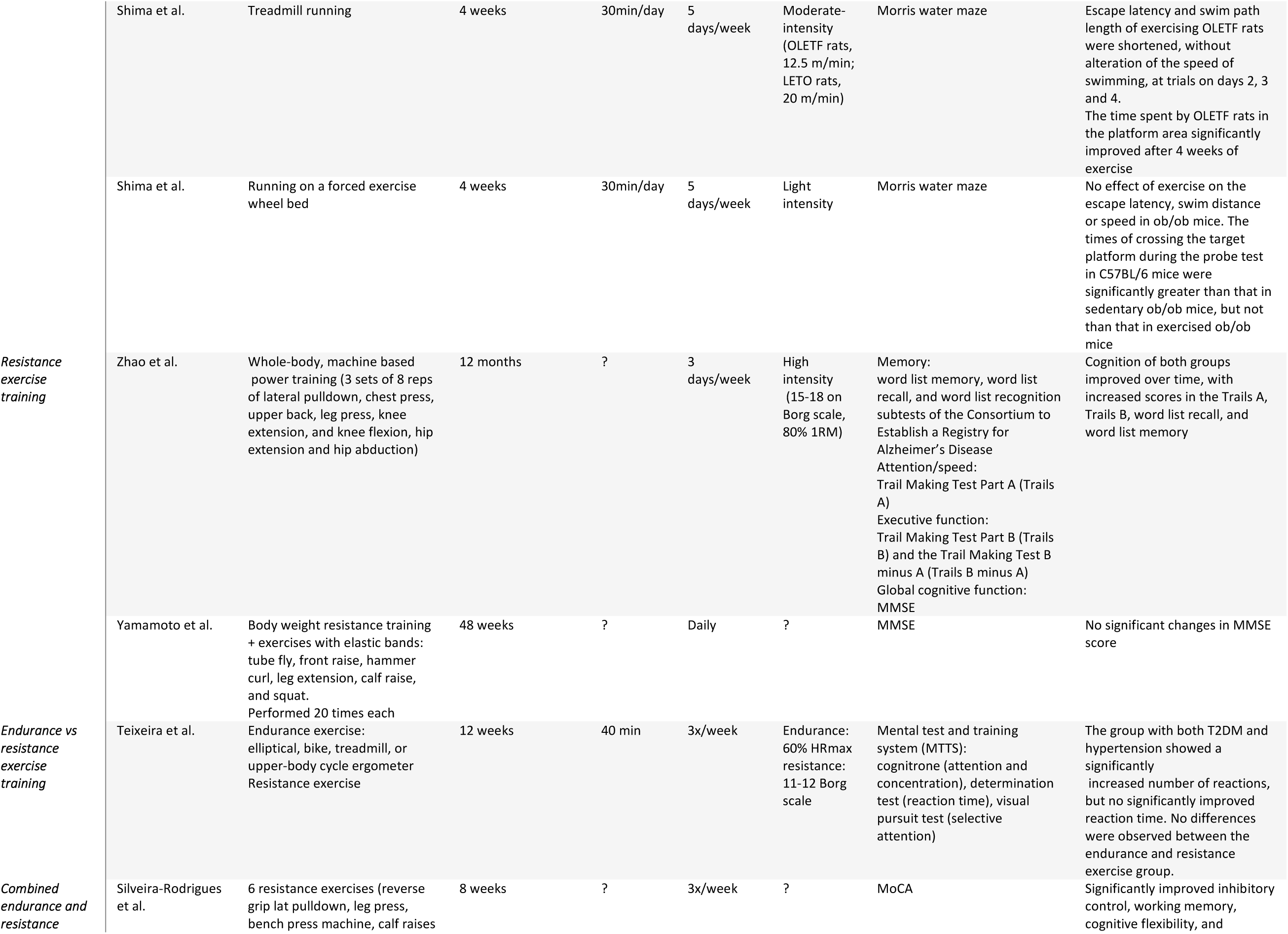
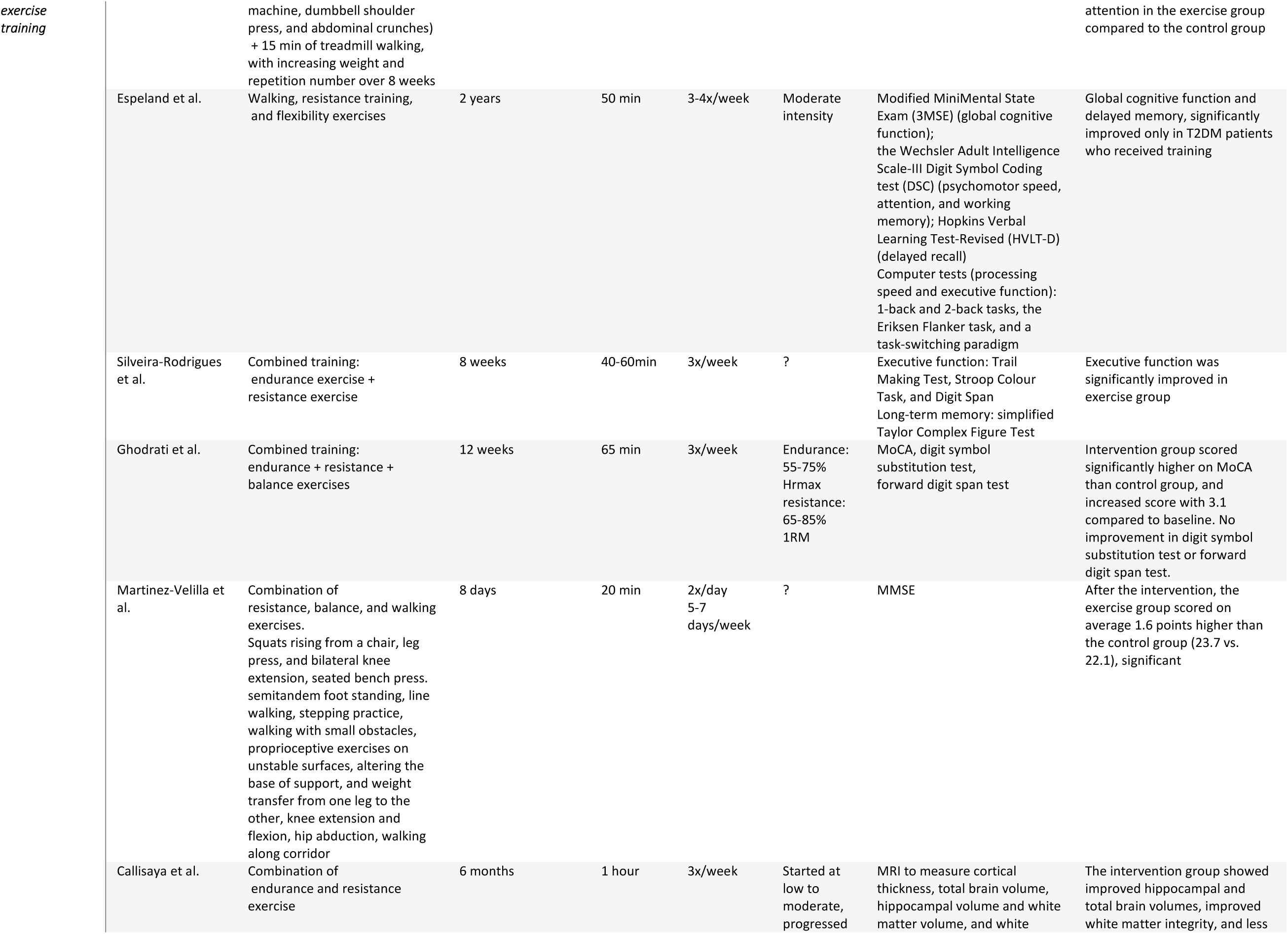
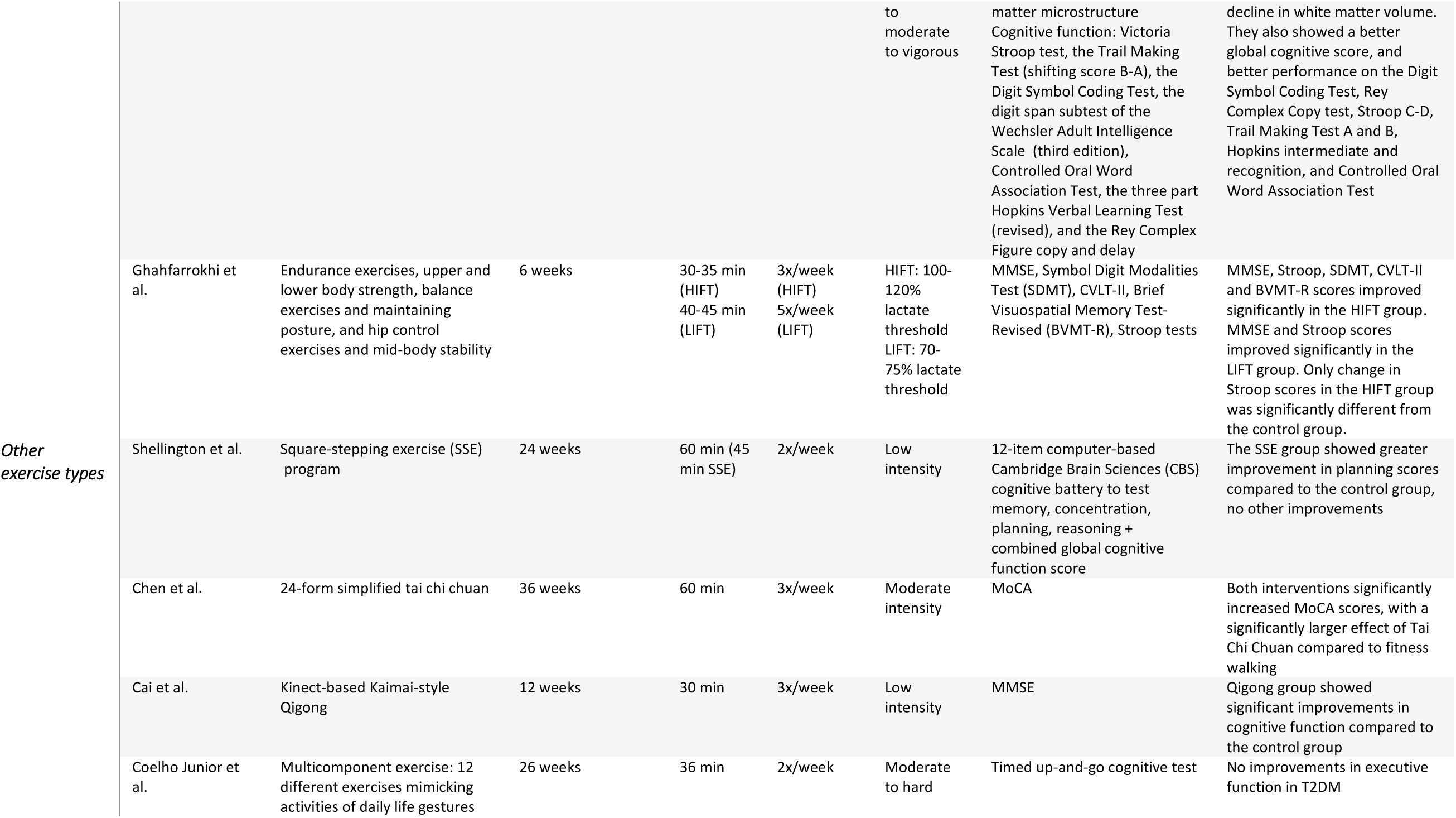
Characteristics of included studies.

### 3.2 Reviewing targets and mediators of the effect of exercise training on the brain in T2DM

To understand why certain exercise trainings cause cognitive improvement in T2DM, and others do not, there is a need for studies investigating the underlying biomedical mechanisms of the effect of exercise on the brain and cognition in T2DM. This knowledge can contribute to the development of pharmacological therapies and exercise programs specifically designed for improving brain health and cognition in T2DM in the most efficient way. The following section discusses such studies, and aims to identify the different targets and mediators involved. Although there is much more extensive literature available on the possible mediators of the effect of exercise on the brain in T2DM, this review will solely focus on the studies in T2DM that have actually demonstrated an involvement of certain mediators.

#### 3.2.1 AD-related pathological markers: amyloid β peptides and hyperphosphorylated tau

Increasing evidence and literature points to converging pathways and pathogenetic processes in T2DM and AD. Here we focus on studies showing effects on amyloid β and tau in T2DM due to exercise training. We refer to extensive reviews regarding T2DM and AD for further reading [24, 104, 105].

##### Leptin

Several studies have indicated that exercise is negatively associated with cerebral amyloid β and hyperphosphorylated tau in T2DM, and contributes to memory maintenance [106, 107]. The aspartyl protease β-site AβPP-cleaving enzyme 1 (BACE1) is responsible for catalysing the rate-limiting step of amyloid β production. It has been shown that the adipocytokine leptin reduces BACE1 activity and expression, thereby limiting the production of amyloid β [108, 109]. This is supported by Rezaei et al. (2023), who suggested leptin as a possible mediator of exercise-induced prevention of memory impairment in T2DM after finding elevated serum and hippocampal levels of this hormone together with decreased hippocampal levels of BACE1, amyloid β and hyperphosphorylated tau after 8 weeks of HIIT training in T2DM rats [110].

##### GSK3β

The maintenance of cognitive function in exercising T2DM rats treated with dexamethasone, demonstrated by De Sousa et al. (2020), was according to the authors presumably mediated by the observed lesser inhibition of the activation of hippocampal IRS-1 and higher concentration of GSK3β phosphorylated on serine-9 (Ser-9) [111]. GSK3β is namely inactivated by Ser-9 phosphorylation [112], which prevents GSK3β activation, and subsequently tau phosphorylation and associated neurotoxic effects [113, 114]. Accordingly, Rezaei et al. (2023) also found decreased hippocampal GSK3β dephosphorylation together with decreased hippocampal levels of amyloid β and phosphorylated tau in HIIT-trained T2DM rats [110]. GSK3β plays an important role in neurodegeneration in T2DM. Both glucolipotoxicity and insulin resistance contribute to GSK3β overactivation, leading to β-catenin phosphorylation and subsequent proteasomal degradation. This results in the inhibition of the expression of reactive oxygen species (ROS) scavenging enzymes, which leads to more oxidative stress and consequently the disruption of mitochondrial structure, function and axonal trafficking. Moreover, GSK3β overactivation also stimulates tau hyperphosphorylation, leading to microtubule destabilisation and thus disruption of axonal mitochondrial trafficking, as well as to a decrease in mitochondrial complex I and thus impaired energy production [114]. By inhibiting the activation of GSK3β through Ser-9 phosphorylation, exercise could thus contribute to the prevention of cognitive decline in T2DM.

##### Adiponectin

In the study of Rezaei et al. (2023), the T2DM rats showed decreased serum and hippocampal levels of insulin and adiponectin at baseline, as well as decreased levels of insulin receptors, adiponectin receptors and AMPK in the hippocampus, and increased hippocampal GSK3β and hyperphosphorylated tau. HIIT training counteracted these findings. Both serum insulin and adiponectin were significantly increased by HIIT, as well as the levels of hippocampal insulin and adiponectin receptors. HIIT training also increased hippocampal AMPK phosphorylation, and decreased hippocampal tau phosphorylation and GSK3β dephosphorylation. The authors suggest that the increased levels of adiponectin can contribute to the preservation of hippocampal volume and function through the stimulation of synaptic plasticity, as it has been shown by Pousti et al. (2018) and Weisz et al. (2017) respectively that adiponectin modulates synaptic plasticity in the hippocampal dentate gyrus and regulates hippocampal synaptic transmission [110, 115, 116]. Another study shows memory and learning impairments together with reduced AMPK phosphorylation and increased GSK3β activation in adiponectin-knockout mice [117], suggesting that the neuroprotective effects of adiponectin are mediated via AMPK phosphorylation and GSK3β inactivation, which is in line with the study of Rezaei et al. (2023). The exercise-induced decreased GSK3β Ser-9 dephosphorylation means that there is a higher ratio of inactive, phosphorylated GSK3β, preventing tau phosphorylation. In addition, the adiponectin-mediated AMPK phosphorylation could also be neuroprotective by inhibiting the inflammatory response of microglia to amyloid β [118].

#### 3.2.2 Brain volume

Several studies have demonstrated that exercise increases hippocampal volume in T2DM patients. One study showed that a higher step count, measured over 7 days, was associated with a larger hippocampal volume in T2DM patients [59]. In addition, one year of endurance training in T2DM patients with normal cognition significantly increased hippocampal volume, and prevented a decline in MMSE and MoCA scores. Hippocampal volume in the training group was significantly increased compared to baseline, as well as compared to the control group [79]. Another study showed a higher hippocampal CA1 and CA3 neuronal density in diabetic Sprague-Dawley (SD) rats after 6 weeks of exercise on a running wheel, probably contributing to the observed improved cognitive function [119]. Similarly, a pilot study in which T2DM patients performed 6 months of a progressive endurance- and resistance-training program showed improved hippocampal and total brain volumes, improved white matter integrity, and less decline in white matter volume in the intervention group compared to the control group [97].

#### 3.2.3 Neuronal survival

##### BDNF

BDNF is one of the most important neurotrophic factors. By binding the TrkB receptor, it regulates and promotes cell survival, neuroplasticity, and neurogenesis in the central nervous system [120, 121]. As already mentioned previously, BDNF plays an important role in the effect of exercise on cognitive function. A study investigating the effect of 4 weeks of swimming training on working, spatial, and recognition memory in diabetic C57BL/6 J mice, shows that higher PA is associated with increased hippocampal and prefrontal BDNF levels, together with an improvement in all memory aspects [80]. Similarly, Jesmin et al. (2022) showed significantly increased hippocampal BDNF levels together with improved memory function in OLETF rats after 4 months of endurance exercise [83]. This shows that also under diabetic conditions, BDNF is an important mediator of exercise effects on cognition.

Another study looked into the effect of 12 weeks of swimming training on BDNF/TrkB signalling and apoptosis in the cerebral cortex of male diabetic C57BL/6JNarl mice. The exercised diabetic mice showed less neural apoptosis compared to the control diabetic mice, accompanied by an increased activity of the BDNF/TrkB signaling pathway, and a decreased activity of the Fas/FasL-mediated and mitochondria-initiated apoptotic pathways. These findings indicate that exercise promotes neuronal survival in diabetic mice through the BDNF pathway [122].

#### 3.2.4 Neuroplasticity and neurogenesis

One study determined the effect of 7 weeks of treadmill exercise on neuroblast differentiation in the subgranular zone of the dentate gyrus (SZDG) in Zucker diabetic fatty (ZDF) rats and Zucker lean control (ZLC) rats. At 23 weeks old, the rats started to exercise for 7 weeks at 12-16m/min. Compared to the non-exercising groups, the neuroblasts of both the exercising diabetic and exercising control rats showed a significant increase in tertiary dendrites. However, the number of neuroblasts only increased in the control rats. This shows that endurance exercise in diabetic rats can stimulate neuroplasticity, but not necessarily neuroproliferation [123]. A similar study tested the effect of 5 weeks of treadmill exercise on cell differentiation and proliferation in the SZDG of the same ZDF rat model. Proliferation was detected by means of the proliferation marker Ki67, and progenitor differentiation into neurons was detected by means of the differentiation marker doublecortin (DCX). In this study, the rats started exercising at 6 weeks old, and ran at 22m/min. Compared to the sedentary ZDF rats, the exercised rats showed a significant increase in Ki67 positive cells and DCX-immunoreactive structures, indicating both increased proliferation and increased differentiation [124]. This confirms that exercise can stimulate neuronal differentiation in the diabetic SDZG, as shown by Hwang et al. (2010) [123]. The fact that, this time, exercise was able to stimulate neuronal proliferation as well, might indicate that exercise has to be performed at a certain intensity rather than a certain volume to achieve this effect.

##### BDNF

Increased hippocampal BDNF levels in combination with increased dendritic spine density on the secondary and tertiary dendrites of dentate granule neurons were found after voluntary wheel running in *db/db* mice [125]. This suggests that BDNF contributes to exercise-induced neuroplasticity in diabetic mouse models.

##### PI3K/Akt and AMPK/SIRT1

It was demonstrated that endurance exercise can upregulate synaptic plasticity-associated proteins in the hippocampus of T2DM rats, presumably via the activation of PI3K/Akt/mTOR and AMPK/SIRT1 signalling pathways and inhibition of the NFκB/NLRP3/IL-1β signalling pathway [126]. Under non-diabetic conditions, the PI3K/Akt/mTOR pathway is activated by insulin, resulting in the formation of dendritic spines and excitatory synapses in hippocampal neurons [127]. In T2DM, insulin resistance prevents this activation, contributing to reduced synaptic plasticity. Exercise thus could, to a certain extent, take over this role of insulin. The aforementioned BDNF also activates the PI3K/Akt pathway, constituting another mediator of the exercise-induced increase in synaptic plasticity-associated proteins [120]. Activation of the AMPK/SIRT1 pathway, on the other hand, contributes to synaptic plasticity by increasing insulin sensitivity and glucose uptake, regulating BDNF expression, promoting neuronal survival, etc. [128–132]. During exercise, AMPK activates SIRT1, after which SIRT1 deacetylates PGC-1α, resulting in its activation. PGC-1α is important for mitochondrial remodelling and biogenesis, contributing to synaptic plasticity [133]. Its activation leads to the upregulation of FNDC5, which in turn crosses the BBB and upregulates hippocampal BDNF expression [120].

Another study also found an increase in the synaptic plasticity proteins synaptophysin (SYN) and N-methyl-D-aspartate receptor (NMDAR) in the prefrontal cortex of diabetic SD rats after 4 weeks of treadmill training. They demonstrated that endurance exercise increased the level of phosphorylated PI3K, suggesting an increased activity of the PI3K/Akt pathway, and increased insulin sensitivity. In addition, they found a slight increase in the phosphorylation and a slight decrease in the acetylation of FOXO1, which is a transcription factor that promotes NF-kB activity and consequently the expression of proinflammatory factors. FOXO1 phosphorylation enables its cytoplasmic retention and thus prevents the transcription of its target inflammatory genes [134]. Since FOXO is a target of Akt, the increased PI3K/Akt activity might be the cause of the increased FOXO1 phosphorylation [135]. Nuclear NF-kB acetylation was found to be decreased as well in the rats that performed endurance exercise, reducing its DNA binding and transcriptional activity [134, 136]. These findings were confirmed by a study showing that 8 weeks of moderate intensity treadmill exercise in diabetic C57BL/ 6 mice improves cognitive dysfunction and increases the release of BDNF through the activation of the SIRT1/PGC-1α/FNDC5/BDNF pathway [84].

#### 3.2.5 Cerebral blood flow

Zhao et al. (2023) investigated the effect of a 2-month moderate-intensity exercise program on brain oxygenation in sedentary older T2DM patients with cognitive impairment. They also looked into the effect of the exercise program on the hemodynamic responses to the Mini-Cog test, which assesses short-term memory and visuo-constructive abilities. The exercise program consisted of a combination of endurance and resistance exercise. They found that the exercise program improved the very low-frequency oscillations (<0.05 Hz) during the Mini-Cog test, as assessed by near-infrared spectroscopy. Since very-low frequency oscillations are crucial for small-vessel function, this indicates that combined exercise has the power to improve cerebral vascular function in T2DM. In addition, the exercise program significantly reduced the oxyhemoglobin (oxyHb) activation in the right superior frontal region during the Mini-Cog test, thereby reducing the overactivation usually observed in T2DM [137].

#### 3.2.6 Glycometabolism

##### Lactate

Since dysregulated peripheral glycometabolism is a hallmark of T2DM, Shima et al. (2017) hypothesised that hippocampal glycometabolic dysfunction might contribute to memory impairment in T2DM [85]. As it has already been demonstrated in healthy animals that moderate exercise enhances memory function and hippocampal glycogen levels [138, 139], they wanted to determine if this was also the case in a T2DM animal model. At baseline, OLETF rats had higher hippocampal levels of glycogen and lower MCT2 expression compared to LETO control rats. They also showed impaired memory. After 4 weeks of treadmill running, the OLETF rats showed even higher hippocampal glycogen levels, in addition to normalised hippocampal MCT2 expression together with improved memory function. Since MCT2 is an important lactate transporter, this suggests that lactate plays a key role in the effect of exercise on glycometabolism and memory in T2DM [85]. In line with this, it was demonstrated that 4 months of treadmill running prevented the progression of cognitive decline in presymptomatic OLETF rats through improved hippocampal MCT2 expression, again confirming the role of lactate in the positive effect of exercise on cognition [83]. Another study by Shima et al. (2023) showed the same effect in an advanced stage T2DM mouse model. Ob/ob mice were subjected to 4 weeks of light-intensity exercise. The exercise program resulted in improved hippocampal MCT2 levels, accompanied by improved hippocampal memory retention [86]. As Soya et al. (2019) mention in their review discussing these findings, the upregulated MCT2 levels enable an increased uptake of L-lactate into neurons, where it can serve as an energy substrate, and act as a signalling molecule to induce the expression of neuroplasticity genes, eventually contributing to the increased memory function observed in these studies [140].

#### 3.2.7 Inflammation

As inflammation is a very broad process, involving many different players, we would like to emphasize again that the pathways and mediators discussed in this review are only the ones that clearly have been shown to be involved in the effect of exercise on the brain in T2DM. For more information about the involvement of inflammation in T2DM, and the effect of exercise on it, we refer to more exhaustive reviews [71, 141].

##### AMPK/SIRT1

The AMPK/SIRT1 pathway is not only important for rescuing cognitive function by increasing BDNF levels, but also by counteracting inflammation. Treadmill exercise in diabetic C57BL/ 6 mice for 8 weeks reduced activation of hippocampal proinflammatory microglia M1, as well as the hippocampal levels of proinflammatory factors IL-1β, IL-6, TNF-α, and increased the expression levels of anti-inflammatory factors IL-10, TGF-β1. This co-occurred with an activation of the SIRT1/NF-κB pathway, which is known to be responsible for counteracting inflammation, suggesting the importance of this pathway in exercise-mediated anti-inflammatory actions in T2DM [84]. A different study also reported a reduction in hippocampal IL-1β and TNF-α, accompanied by cognitive improvement, after T2DM SD rats performed 6 weeks of endurance exercise on a running wheel. However, although we can assume that the AMPK/SIRT1 pathway is involved here as well, the authors did not elaborate on the pathways responsible for the observed reduction in pro-inflammatory factors [119].

#### 3.2.8 Insulin resistance

T2DM-associated insulin resistance is not only present in the periphery, but also in the brain, where it contributes to the cognitive deficits observed in T2DM patients via multiple mechanisms [128, 142–144]. For example, insulin resistance and the consequent hyperinsulinemia have been associated with increased tau hyperphosphorylation, increased deposition together with reduced breakdown of amyloid β, dysfunctional IGF-1 receptors (IGFRs), etc. This all contributes to neurodegeneration and cognitive impairment [142]. Studies have shown that the insulin-sensitising effect of exercise is also manifested in the brain, which gives exercise the power to prevent or decrease these cognitive deficits [145, 146]. One possible way in which this effect of exercise could be mediated, is by the afore-mentioned decrease in TNF-α. TNF-α activates JNK, which is a kinase that causes insulin resistance via serine phosphorylation of IRS-1 [71, 147]. IRS-1 is important for the signal transduction of insulin, ultimately leading to glycogen synthesis and glucose transporter 4 (GLUT-4) translocation. Serine phosphorylation of IRS-1 inhibits this insulin signal transduction and thus results in insulin resistance [148–150]. It could thus be hypothesised that by decreasing TNF-α, exercise could increase insulin sensitivity in the brain.

##### GLP-1

Park et al. (2019) investigated the effect of resistance exercise training on GLP-1R levels in the hypothalamus of OLETF rats. The exercise training comprised 12 weeks of ladder climbing. As a result of the resistance training, the rats showed increased levels of hypothalamic GLP-1R mRNA, protein kinase A (PKA), protein kinase B (PKB), glucose transporter 2 (GLUT-2), and decreased hypothalamic levels of protein kinase C-iota (PKC-ι) [151]. All these findings point in the direction of improved glycaemic control. PKB, also known as Akt, plays an important role in the PI3K/Akt pathway downstream of the IR, as mentioned before in this review [23, 27, 126]. PKA, on the other hand, is essential for the regulation of metabolism and triglyceride storage [152, 153], while PKC-ι is known to cause metabolic abnormalities in T2DM [154, 155]. GLP-1R, the main focus of this study, is a main receptor involved in T2DM. It is responsible for lowering blood glucose levels by, among other things, stimulating insulin secretion and suppressing glucagon secretion [156]. In addition, GLP-1R has been suggested to play a neuroprotective role in diabetes-related neurodegeneration by increasing insulin sensitivity [157]. Moreover, a different study has shown that recombinant human GLP-1 can reduce oxidative stress by activating PKA, which was found to be elevated in the study of Park et al. (2019), and by inhibiting PKC, which was found to be decreased, ultimately reversing diabetic nephropathy [158]. The increase in GLP-1 mRNA found by Park et al. (2019) thus indicates decreased oxidative stress and increased insulin sensitivity in the T2DM rats after resistance exercise training.

#### 3.2.9 Mitochondrial function

Mitochondrial dysfunction is an important contributor to T2DM pathology [159, 160]. Multiple studies have already demonstrated mitochondrial dysfunction in the T2DM brain, where it contributes to neurodegeneration and cognitive dysfunction [161, 162]. It is also known that exercise positively influences mitochondrial function [163–165]. Studies show that exercise can restore mitochondrial function in the muscle of T2DM patients [166–169]. However, little is known about the effect of exercise on mitochondrial function in the brain in T2DM.

##### Insulin sensitivity

One study investigated the effect of endurance exercise training in combination with metformin on mitochondrial function in male C57BL/6J mice with brain insulin resistance induced by a high-fat diet. They observed that the mitochondria in brain regions rich in insulin receptors produced less ATP and showed reduced activity of oxidative enzymes. The mice also showed elevated ROS production and reduced activity of antioxidant enzymes, accompanied by higher rates of mitochondrial fission, and accumulation of damaged mitochondrial proteins. Endurance exercise training together with metformin improved insulin sensitivity. This resulted in reduced ROS emission, less hippocampal mitochondrial fission, less mitochondrial protein oxidation, and increased ATP production in astrocytes and primary cortical neurons. The reduction in ROS emission and increased ATP production were counteracted by intranasal administration of the insulin receptor antagonist S961, proving that these mitochondrial ameliorations were mediated by increased insulin sensitivity [170]. Since metformin was also administered in this study, the observed effects are not fully attributable to exercise, however, it is clear that metformin and exercise have a synergistic effect, and that exercise can certainly be an added value in the treatment of T2DM.

## 4. Discussion

In the systematic part of this review, 24 studies investigating the effect of exercise training on cognition in T2DM were included. Although mixed results were found, most studies (20 out of 24) demonstrated a significant positive effect of exercise training on cognition, which is consistent with findings of previous reviews on this topic [65, 67]. 7 out of 10 studies investigating the effect of endurance exercise training were able to show cognitive improvement, indicating the effectiveness of this training modality. Even endurance exercise as simple as walking showed to already be successful in improving cognition in T2DM. Out of the two studies investigating the effect of resistance exercise on cognition in T2DM, one study failed to show cognitive improvement. This lack of effect could be attributed to the lower intensity or lack of weights in this exercise intervention, although an effect was observed in the sham low-intensity group of the study by Zhao et al. (2022), where also no added weights were used. Additional studies are required to clarify this. Studies comparing endurance exercise training to resistance exercise training found similar results for both exercise forms. Consequently, exercise interventions in T2DM patients aimed at improving cognition do not have to be limited to one type of exercise, and can be adapted to the patient’s capacities. In addition, all included studies investigating the effect of combined (endurance + resistance (+ flexibility/balance)) exercise training were able to show significant cognitive improvements. Being in line with the exercise recommendations of the American College of Sports Medicine for older adults [171], this finding favours combined exercise to reach cognitive improvement in T2DM. Finally, tai chi chuan and Kaimai style Qigong showed to improve cognition in T2DM. The beneficial effect of these exercise forms might be attributed to the fact that they are motorically more complex, and require a more extensive involvement of the mind.

Based on this review, one can thus conclude that both endurance and resistance exercise training, as well as exercise training involving the mind, are effective at improving cognition in T2DM. Combined exercise training is presumably most desirable, although studies comparing combined exercise training to endurance or resistance exercise training alone are lacking. It also seems that a minimal intensity is required in resistance exercise training to elicit significant improvements. However, because no studies directly compare the effect of different exercise training intensities on cognition in T2DM, the optimal exercise intensity for cognitive improvement in T2DM is yet to be determined.

The mechanisms underlying cognitive improvement following exercise in T2DM have been explored in the second, narrative part of this review. Both the AMPK/Sirt1 pathway and PI3K/Akt/mTOR pathway have been identified as key players. The studies included in this review show that exercise-mediated activation of these pathways leads to beneficial brain changes in T2DM, such as increased neuroplasticity, brain volume, synaptic plasticity, dendritic spines, insulin sensitivity, mitochondrial remodelling and biogenesis, and ATP production, as well as decreased inflammation, oxidative stress, and amyloid β and p-tau production. BDNF, lactate, leptin, adiponectin, GSK3β and GLP-1 were identified as the most important factors mediating these changes. Although other factors such as IGF-1 have been suggested to play a role in the positive effect of exercise on cognition [172], no studies in T2DM have directly demonstrated this. Much more research is needed to identify the full spectrum of involved mediators and pathways, investigate interactions between different cell types, etc. In addition, it needs to be considered that the majority of the studies described in this review were conducted in rodents, and are yet to be confirmed in humans.

This review has various limitations that could interfere with the reliable interpretation of its findings. First of all, the fact that the studies included in this review use a variety of cognitive tests, makes it difficult to compare the different studies. Moreover, several studies merely use the MMSE or MoCA to test cognition, which are diagnostic tools designed to screen for MCI, and not to detect (subtle) changes in cognition [87, 88]. There is a need for a standardised cognitive test battery in T2DM to increase the comparability of future studies and draw more reliable conclusions. In addition, the age, medication use, and ethnicity of the patient population varied across studies. Since cognition is heavily influenced by age [173], and medication use [174, 175] and genetics [61] strongly affect the response to exercise, comparability of the included studies may have been limited by these inconsistencies. Regarding the narrative part of this review, an important limitation is that only studies directly demonstrating an involvement of certain mediators in the effect of exercise training on the brain in T2DM have been considered. This means the present review is a non-exhaustive presentation of the involved mediators and pathways. Finally, publication bias might have skewed the results of this review towards a positive effect of exercise on cognition in T2DM.

In order to define an exercise program that is specifically tailored to cognitive improvement in T2DM, future studies should directly compare exercise of different intensities and volumes, as well as different exercise modalities. In addition, as already mentioned, a standardised cognitive test should be used that is most sensitive to picking up exercise-induced cognitive changes in T2DM. Furthermore, medication use, disease stage, age, and genetics should be taken into account.

## 5. Conclusion

Overall, it can be concluded that exercise has a positive influence on cognition and brain structure in T2DM. There are few studies that fail to find a positive correlation between exercise training and cognition in T2DM, which can often be attributed to a small sample size or exercise of too low intensity. From this review, we can assume that resistance and endurance exercise training have similar effects on cognition in T2DM, and that unconventional exercise training involving the mind, such as tai chi, is also capable of inducing cognitive improvements. However, since there is a lack of studies directly comparing different exercise intensities, little can be said about which exercise intensity would elicit the best cognitive results in T2DM. Future studies should look into this in order to make exercise programs for cognitive preservation in T2DM patients as targeted and efficient as possible.

## 6. Declarations

### Author contributions

Jitske Vandersmissen conducted the literature search and wrote the manuscript. Dominique Hansen, Koen Cuypers, and Ilse Dewachter revised the manuscript and provided feedback.

## Data Availability

All data described in this manuscript are available online from the referenced articles.

